# Screening Diabetic Retinopathy Using an Automated Retinal Image Analysis System in Mexico: Independent and Assistive use Cases

**DOI:** 10.1101/2020.07.20.20157859

**Authors:** Alejandro Noriega, Daniela Meizner, Dalia Camacho, Jennifer Enciso, Hugo Quiroz-Mercado, Virgilio Morales-Canton, Abdullah Almaatouq, Alex Pentland

**Affiliations:** MIT Media Laboratory, Massachusetts Institute of Technology, Cambridge, USA; Prosperia Salud, Mexico City, Mexico; Retina Department, Asociación para Evitar la Ceguera en México. Mexico City, Mexico; Engineering Academic Division, ITAM, Mexico City, Mexico; Posgrado de Ciencias Bioquímicas, UNAM, Mexico City, Mexico; Sloan School of Management, Massachusetts Institute of Technology, Cambridge, USA

**Keywords:** Diabetic retinopathy, automated diagnosis, retina, fundus image analysis

## Abstract

**Background:** The automated screening of patients at risk of developing diabetic retinopathy (DR) represents an opportunity to improve their mid-term outcome, and lower the public expenditure associated with direct and indirect costs of common sight-threatening complications of diabetes.

**Objective:** In the present study, we aim at developing and evaluating the performance of an automated deep learning–based system to classify retinal fundus images from international and Mexican patients, as referable and non-referable DR cases. In particular, we study the performance of the automated retina image analysis (ARIA) system under an independent scheme (i.e. only ARIA screening) and two *assistive* schemes (i.e., hybrid ARIA + ophthalmologist screening), using a web-based platform for remote image analysis.

**Methods:** We ran a randomized controlled experiment where 17 ophthalmologists were asked to classify a series of retinal fundus images under three different conditions: 1) screening the fundus image by themselves (*solo*), 2) screening the fundus image after being exposed to the opinion of the ARIA system (*ARIA answer*), and 3) screening the fundus image after being exposed to the opinion of the ARIA system, as well as its level of confidence and an attention map highlighting the most important areas of interest in the image according to the ARIA system (*ARIA explanation*). The ophthalmologists’ opinion in each condition and the opinion of the ARIA system were compared against a *gold standard* generated by consulting and aggregating the opinion of three retina specialists for each fundus image.

**Results:** The ARIA system was able to classify referable vs. non-referable cases with an area under the Receiver Operating Characteristic curve (AUROC), sensitivity, and specificity of 98%, 95.1% and 91.5% respectively, for international patient-cases; and an AUROC, sensitivity, and specificity of 98.3%, 95.2%, and 90% respectively for Mexican patient-cases. The results achieved on Mexican patient-cases outperformed the average performance of the 17 ophthalmologist participants of the study. We also find that the ARIA system can be useful as an assistive tool, as significant sensitivity improvements were observed in the experimental condition where participants were exposed to the answer of the *ARIA system* as a second opinion (93.3%), compared to the sensitivity of the condition where participants assessed the images independently (87.3%).

**Conclusions:** These results demonstrate that both use cases of ARIA systems, *independent* and *assistive*, present a substantial opportunity for Latin American countries like Mexico towards an efficient expansion of monitoring capacity for the early detection of diabetes-related blindness.

## Introduction

Diabetes is one of the most challenging health problems in the world affecting more than 400 million people. Particularly, diabetes threatens the health care systems of low- and middle-income countries where 80% of the world’s diabetic population lives [1,2]. Diabetes is a multifactorial and complex disease with a strong genetic component. In this regard, it has been demonstrated that Hispanic/Latinos have a greater susceptibility to develop type II diabetes (T2D), as well as diabetes-associated complications including renal insufficiency and visual impairment [1–4].

In 2015, there were more than 41 million adults diagnosed with diabetes in Latin America and Caribbean (LAC) countries, making it one of the major causes of premature death and disability in the region [5,6]. Particularly, Mexico ranked sixth in world prevalence of diabetes in 2015, and the 2nd in Latinamerica [7,8]. It is estimated that 26 million adults live with diabetes and only half of them have been diagnosed. Diabetes and its related complications are the first cause of disability and the 3rd cause of death in the country, having a great impact in productivity, life quality and economy [5].

### Evolution and treatment of DR

Among the physiological consequences of an advanced or uncontrolled diabetes, diabetic retinopathy (DR) is the most common microvascular complication and the leading cause of vision loss in working-age adults [9,10]. DR emerges in diabetic patients as a consequence of chronic hyperglycemia that contributes to blood vessels damage in the retina causing fluid leakage, swelling of the surrounding tissue, blood flow obstruction and/or abnormal neovascularization [9,10].

DR progression is slow and gradual, and reversible in its initial stages, however if not treated promptly, evolves to irreversible blindness. The first stages of DR are classified as low and mild non-proliferative DR (NPDR), they are characterized by the presence of at least one microaneurysm and are highly treatable through blood pressure, cholesterol and sugar levels control. Only cases with macular edema might require laser photocoagulation or intravitreal injections. More advanced stages, moderate and severe NPDR, include the presence of hemorrhages, microaneurysms, hard exudates, venous beading and/or intraretinal microvascular abnormalities. At these stages, metabolic control is not sufficient to stop the disease progression and the patient will require invasive treatments like photocoagulation and intravitreal anti-vascular endothelial growth factor (VEGF) agents or corticosteroids. The most advanced stage is proliferative DR (PDR) and is characterized by neovascularization, preretinal hemorrhages, hemorrhage into the vitreous, traction retinal detachments, or macular edema (ME). PDR is treated with a more aggressive laser therapy called scatter or pan-retinal photocoagulation, intravitreal injections, and in some cases, vitreoretinal surgery to remove scar tissue and/or blood from the vitreous cavity, for laser repair of retinal detachments and treatment of macular holes. [10-13]

To prevent the progression of DR to advanced stages, diabetic patients are recommended to have annual or semi-annual retinal screenings beginning at the moment of diabetes diagnosis. However, according to data from the Diabetic Retinopathy Barometer, 27% of people living with diabetes declared that they never discussed eye complications with their doctors or did so only after the onset of complications [14]. Through preventive screenings, 70% of the cases can be captured at initial stages of severity and treated with non-invasive strategies, like metabolic control or photocoagulation [15]. Unfortunately, in most developing countries there is no ophthalmological attention at primary care clinics, and it is only until diabetic patients develop vision attenuation that they are referred to second and third level hospitals to be diagnosed and treated [16]. At this point, significant retinal damage has occurred and even with invasive vitreoretinal surgery or photocoagulation, vision can’t be restored.

The limited access to ophthalmologists and retina specialists at primary care clinics, due to financial and staff limitations at national healthcare institutions, precludes the continuous monitoring of diabetic patients in low and middle income countries like Mexico.

### Challenges of DR screening at a large scale

In Mexico, DR is a leading cause of irreversible blindness among the working-age population [4,13]. Around 30% of the patients diagnosed with diabetes develop DR, and it is estimated that as a consequence of the lack of ophthalmological screening programs and an inadequate control of the glucose levels, this number will elevate up to 72% by 2030 [17]. Although, only 13% of the population living with diabetes have visited an ophthalmologist after their diagnosis [4].

One of the main limitations for the establishment of a systematic eye-screening program is the limited availability of ophthalmologists and their unequal distribution around the country. Based on the 2013 registry of society-affiliated ophthalmologists from the Mexican Society of Ophthalmology, the mean value of ophthalmologists per 100,000 population in the capital and the rest-of-country-areas was around nine and two, respectively [18].

### Automated retinal image analysis (ARIA) for DR screening

In recent years, the juncture between the development of advanced statistical methods, the greater availability of data, and the substantial increase in computing power, has allowed the application of advanced computational methodologies, including artificial intelligence (AI), in diverse medical domains. Among AI use cases for social welfare, AI applications in health domains is one of the fastest-growing sectors, projecting an annual growth above 40% [19]. AI tools have been successfully applied in diagnostics, therapeutics, population health management, administration, and regulation, probing their capacity to augment societies’ ability to provide healthcare, increasing coverage and improving the quality of services provided.

Ultimately, AI applications in healthcare present opportunities to improve quality of life, patients’ prognosis of survival, and optimize human and financial resources [20]. In particular, automated retinal image analysis (ARIA) systems have emerged as a promising solution to massify the early detection of DR at primary care clinics, particularly in resource-constrained developing countries, thereby improving health outcomes and reducing treatment costs.

ARIA systems analyze retinal fundus images by applying techniques like deep learning (DL) to classify diabetic patients in a) cases without retinal lesions associated to DR (*non-referable* output) and b) cases that need to undergo examination by an ophthalmologist to confirm diagnosis and define treatment (*referable* output) [21–25]. As of today, various analysis systems have been developed and implemented on the market in European countries, Canada and the United States. Though, very few have been tested in LAC countries in order to evaluate their performance and usability in different contexts [26], considering patients ethnicity, training of the healthcare personnel, community openness to new technologies, and hospital resources as players for their successful implementation.

### Aims and key findings of the study

The present work evaluates the performance of a DL-based ARIA system that classifies retinal fundus images in non-referable or referable, as well as the potential benefits of its use as an assistive tool for ophthalmic doctors. We report on a randomized controlled trial where the performance of the ARIA system was compared to the accuracy of 17 ophthalmologists of one of the most reputed ophthalmic hospitals in Mexico. In particular, we assessed performance of ophthalmologists in three experimental conditions: one *independent* condition, where the ophthalmologists assess the images independently from the ARIA system, and two *assistive* conditions, in which ophthalmologists can observe and be influenced by the opinion, confidence, and/or attention heatmap of the ARIA system.

#### Key findings

The ARIA system developed using DL strategy was able to classify referable vs. non-referable cases with an area under the Receiver Operating Characteristic curve (AUROC), sensitivity, and specificity of 98%, 95.1% and 91.5% respectively, for international patient-cases; and an AUROC, sensitivity, and specificity of 98.3%, 95.2%, 90% respectively for Mexican patient-cases. The results achieved on Mexican patient-cases outperformed the average performance of the 17 ophthalmologist participants of the study. Moreover, we find that the ARIA system can be useful as an assistive tool, as we found significant improvement in the specificity in the experimental condition where participants were able to consider the answer of the *ARIA system* as a second opinion (87.3%), compared to the specificity of the condition where participants assessed the images independently (93.3%).

Hence, the present study demonstrates a high potential value of the use of ARIA systems, in both independent and assistive schemes, towards effectively massifying the early detection of DR in developing countries like Mexico.

## Methods

### ARIA system design

The ARIA system consisted of an image preprocessing module, and an image analysis module that returns a binary referable and non-referable DR classification, the level of confidence of that classification, and an attention map that showed, pixel-wise, the indicative features for referable DR according to the model (see Figure 1). The models constituting the ARIA system were implemented using the Keras library with the Tensorflow backend [27] in Python 3.5 [28].

**Figure 1.**
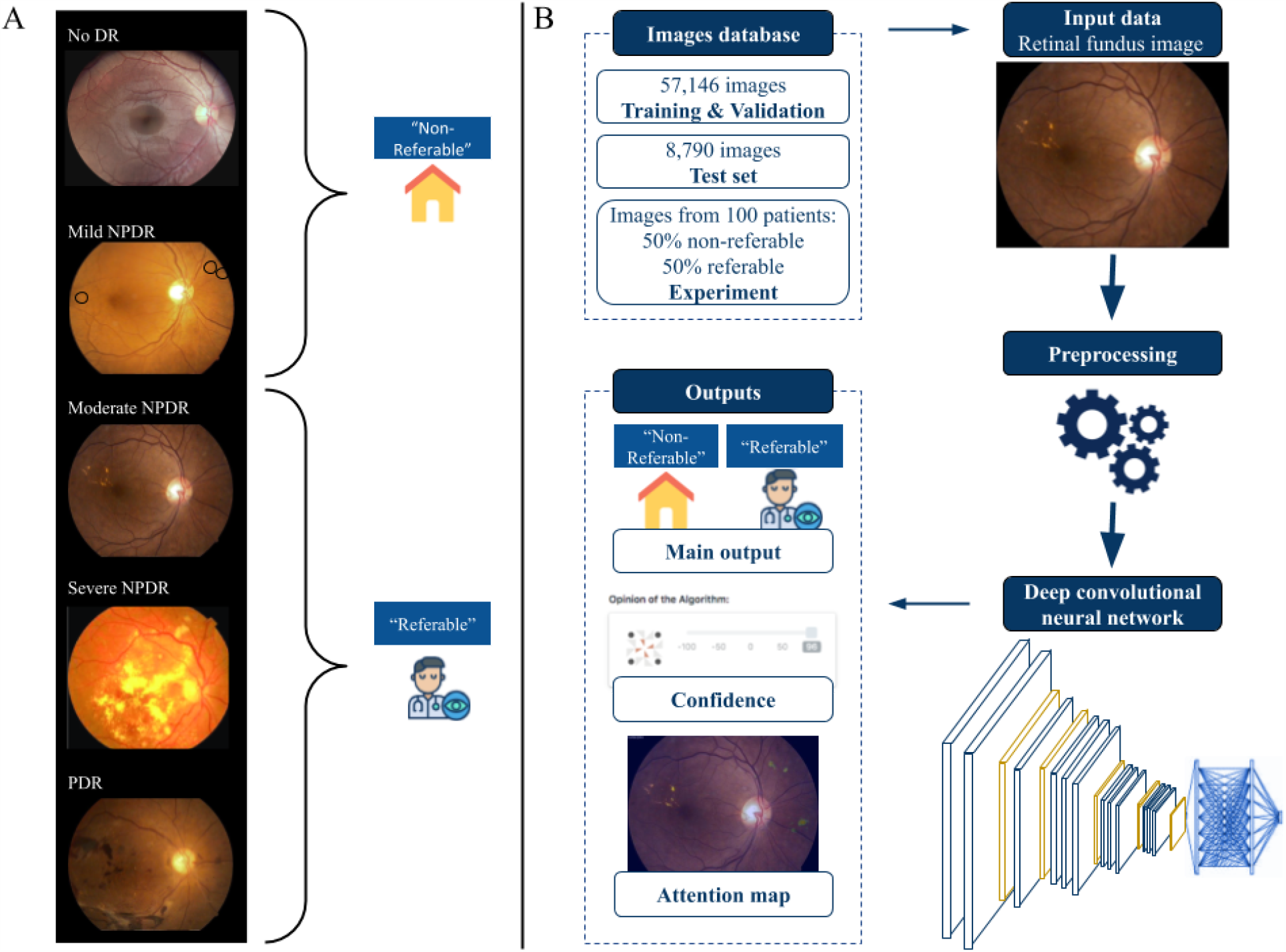
DL-based ARIA system. A) Example of classified retinal fundus images according to the DR severity scale used for the training data. B) Flow chart describing the design of the ARIA system; the data used for training, validation and test; and the algorithm’s outputs.

Images from all datasets were annotated by ophthalmic specialists for 5-class identification according to the International Clinical Diabetic Retinopathy Severity Scales, and subsequently labeled as non-referable and referable DR [30]. Table 1 describes the classification, and Figure 1A provides a graphical example. The *gold standard* classification used for the experimental phase of the study was provided by three retina specialists, as described in the following subsections.

**Table 1.**
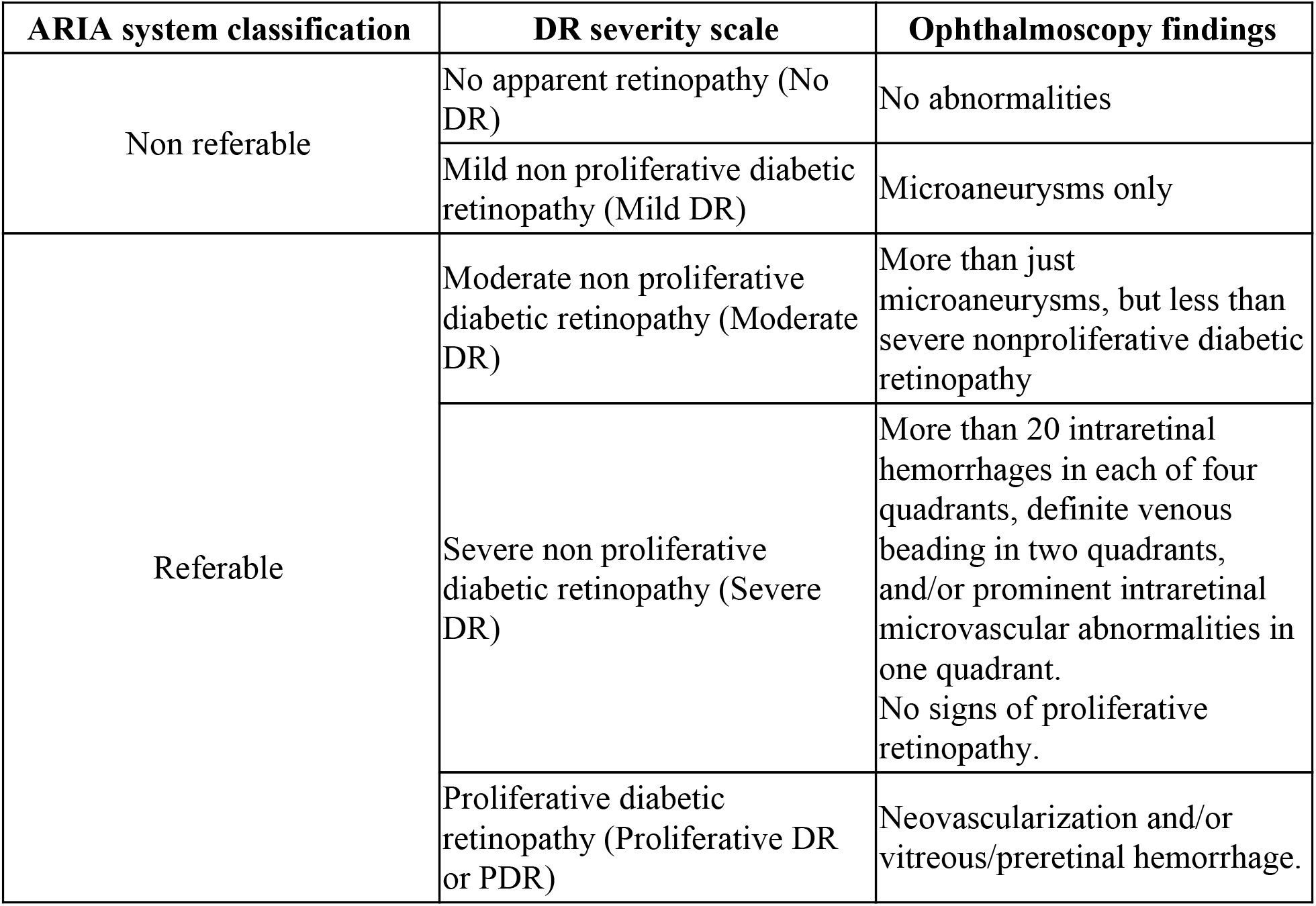
International clinical diabetic retinopathy severity scales and their classification for the ARIA system [29].

#### Preprocessing

Before training or classifying images a preprocessing procedure was applied. The procedure consisted of cropping the background to eliminate non-informative areas, padding to guarantee consistent squared image ratios, resizing the image to 224×224 pixels, and normalizing pixel values to the range 0 to 1.

#### Image classification

The image classification model used was a deep convolutional neural network [31,32], composed of 16 convolutional layers, a dense layer of 1,024 neurons, two dropout layers to avoid overfitting, and a binary classification layer of a single unit with sigmoid activation. Hence, the model output is a value between 0 and 1, that may be interpreted as the confidence of the model regarding a referable DR classification. Lastly, a threshold of 0.5 was used to classify non-referable and referable DR.

#### Attention heatmaps

We developed attention heatmaps to show the importance that each pixel had towards a referable DR diagnosis according to the model. These heatmaps were obtained by applying the layer-wise relevance propagation (LRP) method with an alpha-beta rule [33,34]. In essence, the LRP method redistributes the output value throughout the layers until the input layer (input image) is reached. Figure 2 shows a couple of examples of fundus images and their heatmaps.

**Figure 2.**
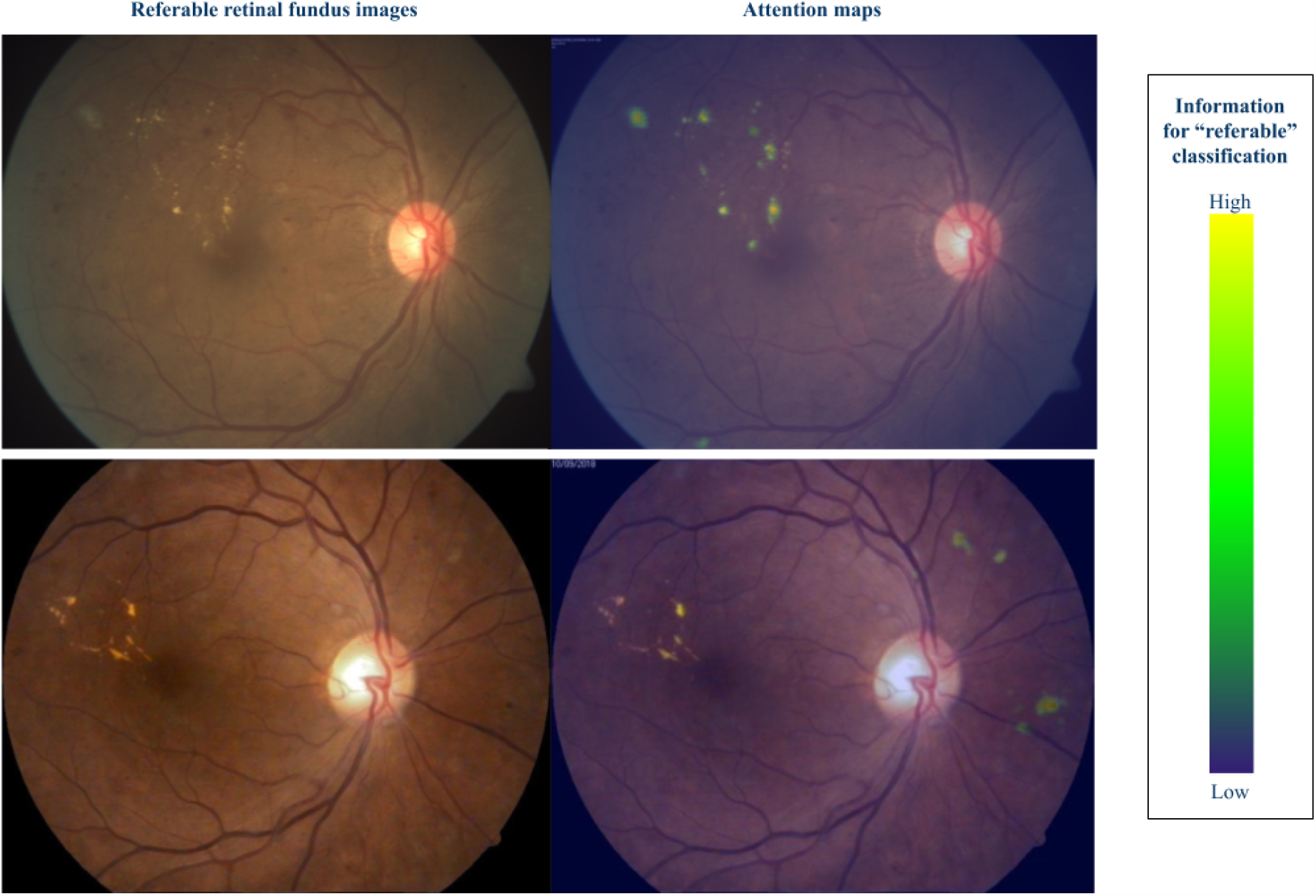
Attention heatmaps for two *referable* images. Green and yellow colors indicate regions in the image that provide information to the algorithm to classify the image as *referable*.

### Study populations

Seventeen ophthalmologists from the Mexican ophthalmic hospital participated in the experimental study, and three retina specialists from the same institution participated in the generation of the *gold standard*. The 17 ophthalmologists evaluated 45 good-quality retinal fundus images each, from 100 Mexican patients, where 50% had non-referable DR levels, and 50% had referable DR levels. The ophthalmologists were retina specialization resident students, with the following distribution: 3 in their second year, 13 in their third year and two in their fourth year of residency.

### Experimental design

We conducted a randomized controlled experiment to assess the performance of the ARIA system in comparison with ophthalmic doctors of the Mexican hospital, as well as to evaluate the potential benefits of using the system as an assistive tool for doctors. To achieve this we developed a web-based experiment platform where ophthalmologists evaluated fundus retinal images under three different conditions—*solo, ARIA answer*, and *ARIA explanation—*described below. Figure 3 displays the main screens of the web platform.

**Figure 3.**
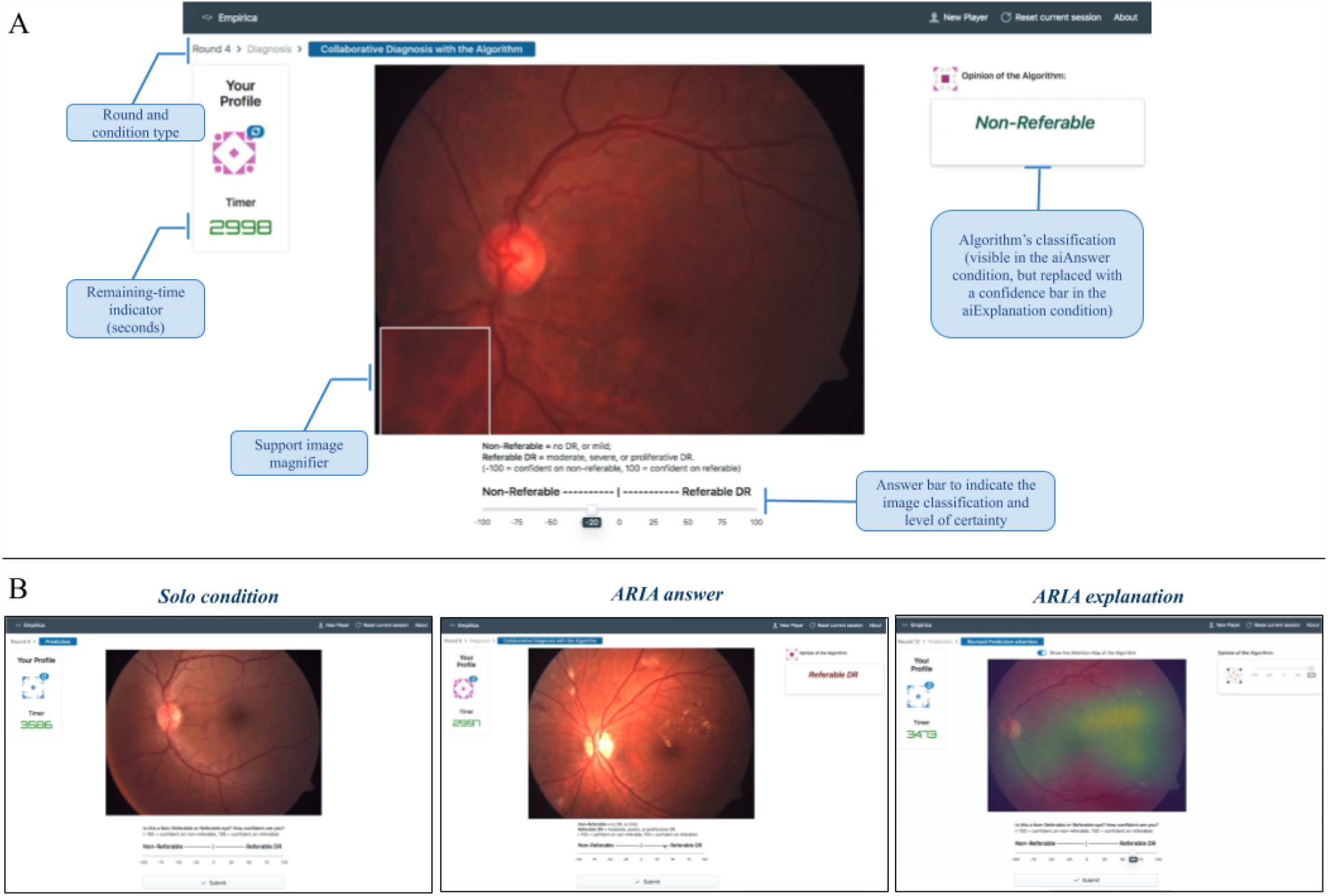
Web-platform design for patient-case classification. A) Visual indicators and components of the classification window, B) visualization of the three experimental conditions.

#### Gold standard and image quality

In order to generate a *gold standard* all fundus images used in the experiment were graded by three retina specialists of the ophthalmic hospital, and a majority rule was used, i.e. if there was a disagreement in the non-referable/referable label, the label selected by two out of three experts was considered the *gold standard*. Image grading was done in the web-based platform. The retina specialists also graded the image quality, and images graded as having bad quality were not considered for the experiment. From the remaining, images from 50 patients with referable DR, and 50 with non-referable DR, were selected at random to be used for the study.

#### Experimental conditions

The experiment followed a within-subjects design, where each ophthalmologist evaluated 45 randomly selected fundus images, 15 for each of the three treatment conditions: *solo, ARIA answer* and *ARIA explanation*. The ophthalmologists were first asked to evaluate 15 fundus retinal images in the *solo* condition, followed by 30 images that randomly alternate between the *ARIA answer* and the *ARIA explanation* conditions. In the *solo* condition, participants responded to the task in isolation, without any exposure to the ARIA system. In contrast, in the *ARIA answer* condition, participants were exposed to the binary answer of the ARIA system (i.e., non-referable or referable), as a second opinion, and then asked to submit their post-exposure answer. The *ARIA explanation* condition was identical to the *ARIA answer* condition, with the exception that participants were shown not only the binary answer of the ARIA system, but also its level of confidence and attention heatmap.

The time for analysis and decision submission per image was limited to 3500 seconds (although the vast majority took less than a minute). Finally, after completing all the classification tasks, the ophthalmologists were asked to submit an optional feedback survey about their experience.

The study was reviewed and approved by the Committee on the Use of Humans as Experimental Subjects at the Massachusetts Institute of Technology, and all participants provided explicit consent prior to their participation.

## Results

### ARIA performance

The ARIA system was first tested in the large dataset of international cases. It there achieved an out-of-sample area under the Receiver Operating Characteristic curve (AUROC) of 98%. In particular, using a given acceptance threshold, the ARIA system achieved a sensitivity of 95.1% and a specificity of 91.5%. Most importantly, the ARIA system was also rather accurate in the patient cases from the Mexican ophthalmic hospital, where it had an AUROC of 98.3%, a sensitivity of 95.2%, and specificity of 90% (see Figure 4).

**Figure 4.**
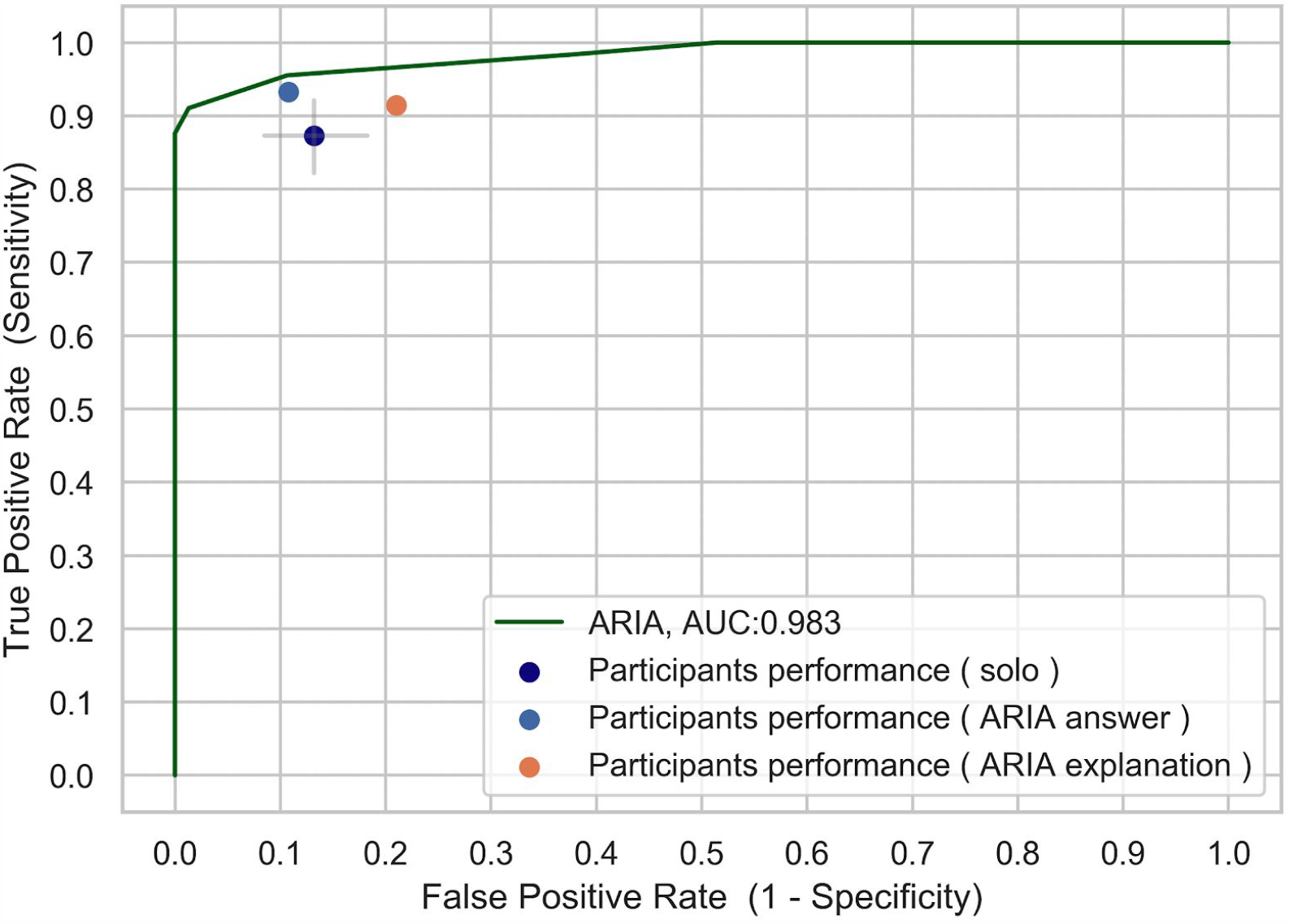
ROC curve of the ARIA system compared to the ophthalmologist’s accuracy under the three experimental conditions (*solo, ARIA answer* and *ARIA explanation*). Grey lines indicate 95% confidence intervals for the *solo* condition.

### Participants’ performance

Figure 4 shows the sensitivity and false positive rate (*1 - specificity*) for each condition—*solo, ARIA answer*, and *ARIA explanation*—and compares them with the ROC curve of the ARIA system. The average sensitivity in the *solo* condition across the seventeen participants was 87.3%, whereas the average specificity was 86.8%. In comparison, the average sensitivity and specificity across participants for the *ARIA Answer* condition were 93.3% and 89.3%, and the average sensitivity and specificity across participants for the *ARIA Explanation* condition were 91.5% and 79%. Hence, we find that the ARIA system was more accurate than the average of participants under any of the three exposure conditions. In particular, the ARIA system could increase sensitivity from 87.3% to 93.3% (vertical movement between the dark blue dot and the green line in Figure 4), while maintaining participants’ specificity of 86.8% constant; or increase specificity to 100% while maintaining participants’ average sensitivity of 87.3% constant (horizontal movement from the dark blue dot leftwards to the green line in Figure 4).

Most interestingly, Figure 4 shows that exposure to the ARIA system was able to improve the performance of human experts, particularly in the *ARIA answer* condition, which significantly improved the sensitivity and specificity compared to the *solo* condition (distance between dark blue and light blue dots in Figure 4). However, performance in the *ARIA explanation* condition had mixed results, improving sensitivity, but lowering the specificity (distance between dark blue and orange dots in Figure 4).

Figure 5 provides more detail on the effect that exposure to information of the ARIA system had on the performance of ophthalmologists. In particular it shows the accuracy (% of correct answers) of the 17 experts consistently improved in the *ARIA answer* condition, shifting the distribution upwards, and decreasing the variance across participants. For example, while only two participants had a perfect score in the *solo* condition, up to 6 participants had a perfect score in the *ARIA answer* condition. However, the *ARIA explanation* condition had mixed beneficial and detrimental effects on participants’ accuracy, and increased the variance of performance across participants compared to the *solo* condition.

**Figure 5.**
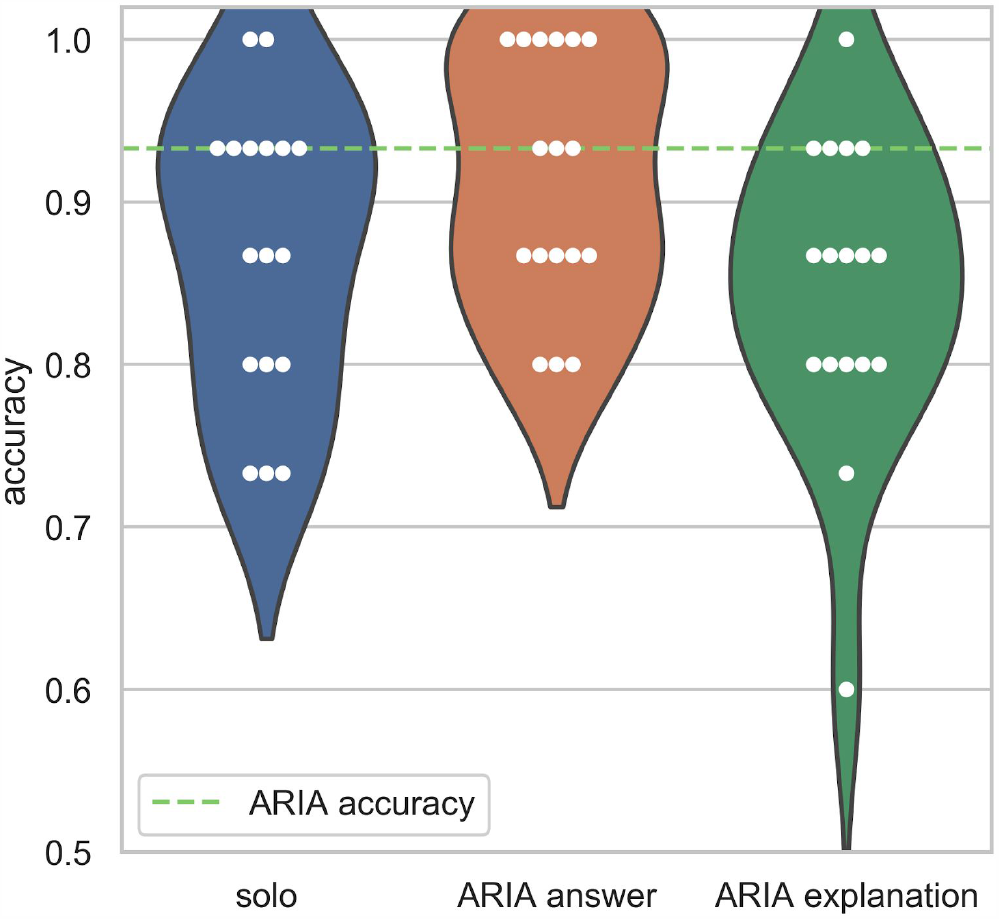
Influence of the ARIA system on the ophthalmologists decision. A) Ophthalmologists performance and B) opinion shift after exposure to the AI outputs.

## Discussion

The number of people living with diabetes is projected to rise around 50% for 2045, reaching a 700 million population worldwide [7,35]. This means that approximately 230 million patients will develop DR and will require routinary eye screenings to be treated on time and prevent vision loss. Just in Mexico, the prevention of DR in patients living with diabetes would implicate savings for up to $10 million dollars to the three main public institutions of the national health care system [36]. The development or ARIA systems represent a possible solution to the increasing demand of eye screenings in healthcare systems, particularly in limited-resource settings.

### Principal results

The DL-based ARIA system presented in this work was evaluated with a subset of retinal images from international patient-cases and an image set of patient cases from a Mexican ophthalmic hospital. In both datasets, the ARIA system outperformed the average sensitivity and specificity of 17 ophthalmology residents of retina specialty. The reached sensitivities (95.1% and 95.2% for the international and mexican datasets, respectively) are comparable to those reported for other seven automated DR screening systems reported in a systematic review, whose sensitivity values were between 87% and 95% [36]. On the other hand, the specificities reached by our ARIA system (91.5% and 90% for the international and mexican datasets, respectively) were higher than the reported average in [36], whose specificity values were between 49% and 69%.

There are now many works demonstrating good overall performance of AI-based ARIA systems for the detection of DR, however sensitivity and specificity are not the only parameters that guarantee a successful implementation in real-life [37,38]. On this line, the evaluation of the ARIA system included two assistive or human-AI hybrid decision schemes, considering that in real-life applications the results of an automated system are reviewed and confirmed by healthcare professionals to choose the most adequate therapeutic protocol for each patient. In these assistive evaluations, we confirmed the existence of significant synergies derived from the interaction of the human and AI dyads. AI’s output exerted a strong influence on the opinion of human participants, however its effect on ophthalmologist’s overall precision depended on the format of ARIA system’s output. A simplified output (i.e. non-referable or referable classification) positively influenced humans’ sensitivity and specificity, while a complex output (i.e. confidence bar and attention map) partially improved human’s decisions, increasing their sensitivity but also the incidence of false positive classifications. These results are coherent with some of the ophthalmologist’s feedback submitted after the classification tasks, where some of them expressed that even when attention heatmaps were useful, the bar showing the confidence of the ARIA system was confusing.

## Limitations

Further pilot studies with a larger number of patients and ophthalmologists will be useful to confirm the ARIA system accuracy. Also, further studies might include direct ophthalmoscopy by retina specialists as the *gold standard*, in order to avoid error related to image quality.

Additional experiments with alternative platform designs might be useful to generate a suitable screening tool that optimizes patient evaluation and referral in three stages. In the first stage, an ARIA system might be useful to identify patients with a higher probability of developing DR. In a second filter, ophthalmologists would be able to evaluate the retinal images of high-risk patients, in combination with the ARIA system output to make a first decision about the disease stage, treatment and finally, refer only patients with an advanced disease to retina specialists.

## Conclusions

Given the short supply of ophthalmologists in the public health system in Mexico, the results of the present study demonstrates a substantial opportunity for Latin American countries like Mexico towards an efficient massification of monitoring systems for early detection of diabetes-related blindness.

The web-based platform developed for this study was designed for the implementation of the ARIA system as an automatic screening tool and as a telemedicine platform useful to confirm or reject the ARIA system’s output with assessment of an ophthalmologist or retina specialist. The platform was useful for the present study, and can be easily adapted for further studies that include the recopilation of additional information about other eye diseases detectable by image analysis (i.e. glaucoma, age-related macular degeneration or coat disease).

With these results we conclude that the proposed ARIA system is useful in an independent and assistive condition, and can be useful to improve DR diagnosis, as well as other ophthalmic diseases in the long-term. However special attention in the design of a careful and explanatory platform, is required for a successful deployment.

## Data Availability

The deep learning model was trained with an open-source database available at Kaggle:Diabetic Retinopathy Detection.

https://www.kaggle.com/c/diabetic-retinopathy-detection

## Acknowledgments

The authors gratefully thank the retina specialists and the ophthalmologists from the APEC hospital involved in this study for their evaluations of the retina fundus images.

## Authors’ Contributions

AN conceived and designed the experiments, analyzed data and contributed to the discussion and review of the paper. DC was responsible for the models’ training, performed the experiments, analyzed data and contributed to the discussion of the paper. DM contributed with the experimental design, image classification and discussion of the paper. JE contributed with data analysis, paper writing and discussion. HQM, VMC, AA and AP contributed to various aspects of the paper including experimental design, machine learning strategies, medical feedback, image evaluations and paper discussion.

## Abbreviations

AI: artificial intelligence
ARIA: automated retinal image analysis
AUROC: area under the Receiver Operating Characteristic curve
DL: deep learning
DR: diabetic retinopathy
FPR: false positive rate
LRP: layer-wise relevance propagation
ROC: receiver operating characteristic
TPR: true positive rate

